# Prediction of serious complications in patients with cancer and pulmonary thromboembolism: validation of the EPIPHANY Index in a prospective cohort of patients from the PERSEO Study

**DOI:** 10.1101/2022.03.28.22272682

**Authors:** Manuel Sánchez-Cánovas, Paula Jimenez-Fonseca, David Fernández Garay, Mónica Cejuela Solís, Diego Casado Elía, Eva Coma Salvans, Irma de la Haba Vacas, David Gómez Sánchez, Ana Fernández Montés, Roberto Morales Giménez, Mercedes Biosca Gómez de Tejada, Virginia Arrazubi Arrula, Silvia Sequero López, Remedios Otero Candelera, Cristina Sánchez Cendra, Marina Justo de la Peña, Diana Moreno Muñoz, Mayra Orillo Sarmiento, Eva Martínez de Castro, Ignacio García Escobar, Alejandro Bernal Vidal, Laura Ortega Moran, Andrés J. Muñoz Martín, Rodrigo Sánchez Bayona, María José Martínez Ortiz, Francisco Ayala de la Peña, Vicente Vicente, Alberto Carmona-Bayonas

## Abstract

**Introduction:** There is currently no validated score capable of classifying cancer-associated pulmonary embolism (PE) in its full spectrum of severity. This study has validated the EPIHANY Index, a new tool to predict serious complications in cancer patients with suspected or unsuspected PE.

**Method:** The PERSEO Study prospectively recruited individuals with PE and cancer from 22 Spanish hospitals. The estimation of the relative frequency θ of complications based on the EPIPHANY Index categories was made using the Bayesian alternative for the binomial test.

**Results:** Nine hundred patients diagnosed with PE between 2017/2020 were recruited. The rate of serious complications at 15 days was 11.8%, 95% highest density interval [HDI], 9.8-14.1%. Of the EPIPHANY low-risk patients, 2.4% (95% HDI, 0.8-4.6%) had serious complications, as did 5.5% (95% HDI, 2.9-8.7%) of the moderate-risk participants and 21.0% (95% HDI, 17.0-24.0%) of those with high-risk episodes. The EPIPHANY Index correlated with overall survival. Both the EPIPHANY Index and the Hestia criteria exhibited greater negative predictive value and a lower negative likelihood ratio than the remaining models. The incidence of bleeding at 6 months was 6.2% (95% HDI, 2.9-9.5%) in low/moderate-risk vs 12.7% (95% HDI, 10.1-15.4%) in high-risk (p-value=0.037) episodes. Of the outpatients, complications at 15 days were recorded in 2.1% (95% HDI, 0.7-4.0%) of the cases with EPIPHANY low/intermediate-risk vs 5.3% (95% HDI, 1.7-11.8%) in high-risk cases.

**Conclusion:** We have validated the EPIPHANY Index in patients with incidental or symptomatic cancer-related PE. This model can contribute to standardize decision-making in a scenario lacking quality evidence.

**Summary:** We have validated the EPIPHANY Index in patients with acute, incidental, or symptomatic cancer-related PE. This predictive model of complications can contribute to standardize decision-making in a scenario lacking quality evidence.

## INTRODUCTION

Pulmonary embolism (PE) has classically been deemed one of the most common and dire complications in patients with cancer [1]. Clear proof of this, the 30-day mortality rate due to acute, symptomatic PE was 21.3% (95% confidence interval [CI], 18.2-24.8) in the EPIPHANY Study [2], often requiring intensive hospital management. In contrast, in recent years, computed tomography pulmonary angiogram (CTPA) and multidetector CT have enhanced the accuracy of PE diagnoses [3,4], with increased detection of unsuspected events. The upsurge of this modality has utterly changed the prognostic scenario of some of these PEs, which is reflected in the epidemiological panorama. In a Danish populational registry, 16% of the cases had cancer and, while incidence of PE increased from 2004 to 2014, the episodes detected were gradually less lethal [5]. More specifically in the oncological population, the pooled frequency of incidental events discovered during cancer staging was 3.2% in a recent meta-analysis [6], exceeding the estimation made one decade earlier [7]. With these data, incidental PE accounts for more than 50% of all episodes of cancer-related PE [4,8]. Given that the prevalence of PE in autopsy series is calculated to be 15-30% [9], incidental diagnoses can be expected to increase, as technology improves.

Ironically, this trend affects decision making in the oncologic population, inasmuch as there is currently no validated method of classification that facilitates choosing episodes with a good prognosis for ambulatory management [2,10–12]. Inarguably, averting hospitalizations would lower costs, avoid iatrogenesis, and improve quality of life [13]. Nonetheless, outpatient management of cancer-related PE is not based on quality evidence, and there is a glaring dearth of prospective data [14]. To date, clinical trials of home management have selected participants using decision rules, such as the Hestia criteria or the like, contraindicating lowering the level of support in patients with any exclusion criterion [15–17]. These exclusion criteria are based on the combination of altered vital signs and factors that point toward a high risk of bleeding or other contraindications to receiving treatment in the home. Nevertheless, there are pockets of uncertainty, given the oncological population’s greater risk of rethrombosis or major bleeding [18,19], the greater multifariousness of PE, and scant experience [12,16,20].

Presently, there is no validated score capable of classifying unsuspected, cancer-related PE. Surprisingly, the scores developed specifically for individuals with cancer (e.g., RIETE or POMPE-C) are not suitable for stratifying incidental PE nor do they predict key outcomes, such as bleeding [2,21,22]. Meanwhile, validated scores for acute, symptomatic PE in the general population (e.g., simplified PESI) fail to capture the heterogeneity of the patient with cancer [2,19,23–25].

The EPIPHANY Index was designed as a modified extension of the Hestia clinical decision rule to pragmatically select patients with low-risk, cancer-related PE [26]. In contrast with other proposals, the EPIPHANY Index is applicable across the entire gamut of PE severity (suspected or unsuspected). A meta-analysis comparing the accuracy of clinical decision rules found that higher sensitivity is obtained with the Hestia criteria and the EPIPHANY Index [27]; however, validations of both sets of criteria in patients with cancer are meager [28].

Against this background, the PERSEO (“*Pulmonary Embolism Risk Stratification & End results in Oncology*”) Study pursues the prospective, multicenter validation of the EPIPHANY Index and Hestia criteria in a broad sample of patients with cancer, with a view to their possible appropriateness in decision making.

## METHOD

### Patients and study design

PERSEO prospectively and consecutively recruited patients with PE and cancer from 22 Spanish tertiary hospitals. Eligibility criteria comprised adults (≥18 years of age) with an active solid neoplasm or anti-tumor treatment at the time of the thrombotic event or in the previous 30 days. All the participants had to have a diagnosis of PE confirmed by an objective imaging technique (CTPA, high probability scintigraphy, or CT scheduled to assess tumor response or for other reasons). In the case of multiple events, only one episode of PE per subject (the first one) was allowed. Subjects were recruited by medical oncologists in outpatient clinics, the Emergency Department, or during hospitalization. No recommendation was made regarding management on the basis of any given clinical decision rule or specific treatment approach, and the cases were managed as per regular practice. The investigators were not blind to outcomes, albeit nor were they aware of the EPIPHANY Index during management. To detect complications, all patients still alive were followed for a minimum of 30 days. In the case of participants treated on an ambulatory basis, follow up was conducted by phone and in outpatient clinics.

The study was approved by the Ethics Committees of all the participating centers and was conducted in accordance with the requirements put forth in the international guidelines regarding epidemiological studies [29], as well as the Declaration of Helsinki and its subsequent revisions. All participants signed an informed consent form.

### Objective

The main objective was to validate the EPIPHANY Index and Hestia criteria in subjects with cancer. Secondary objectives consisted of (1) comparing predictive parameters against other models; (2) assessing the prognostic effect of PE presentation; (3) analyzing bleeding events, rethrombosis, and mortality, and studying the evolution of participants treated on an ambulatory basis.

### Variables

To perform this validation, serious complications within 15 days were factored in [8]. This endpoint includes the development of any of the following events: hypotension (systolic blood pressure <90 mmHg), acute respiratory insufficiency, fibrinolysis, major bleeding (intracranial, intraspinal, intraocular, retroperitoneal, or pericardial, associated with decreased hemoglobin by at least 2 g/dl or requiring the transfusion of two units of red blood cells), acute heart failure secondary to right ventricle failure, acute kidney failure, admission into the Intensive Care Unit, need of cardiopulmonary resuscitation, non-invasive mechanical ventilation, oro-tracheal intubation or death. To be deemed outcomes, these events must have occurred after the objective diagnosis of PE. Should they have occurred before then (e.g., at the time of debut), the events were coded as predictors.

The Hestia criteria [15] (systolic blood pressure <100mmHg, arterial oxygen saturation <90%, respiratory rate ≥30 breaths per minute, pulse ≥110 beats per minute, sudden or progressive dyspnea, other serious complications, constituting admission criteria in and of themselves) were adapted to the oncological population, including: clinically relevant bleeding, high risk of bleeding, or platelets <50,000mm^-3^. The EPIPHANY Index [30] (**Figure 1**) incorporates these criteria together with other particular characteristics of individuals with cancer, such as the Eastern Cooperative Group Performance Status (ECOG-PS), evaluation of tumor response prior to or during the study of the PE using RECIST v1.1 criteria, previous primary tumor resection, oxygen saturation, and the presence or absence of PE-specific symptoms (acute or progressive dyspnea, chest pain, or syncope). The EPIPHANY index is accessible via the web: https://www.prognostictools.es/epiphany/inicio.aspx. Patient and cancer baseline characteristics were attained from the clinical history and from the interview at the time of the PE. Missing for these predictors were not allowed. Overall survival (OS) was estimated from the date of the PE until demise due to any cause, censoring all subjects without any event at last follow up.

**Figure 1.**
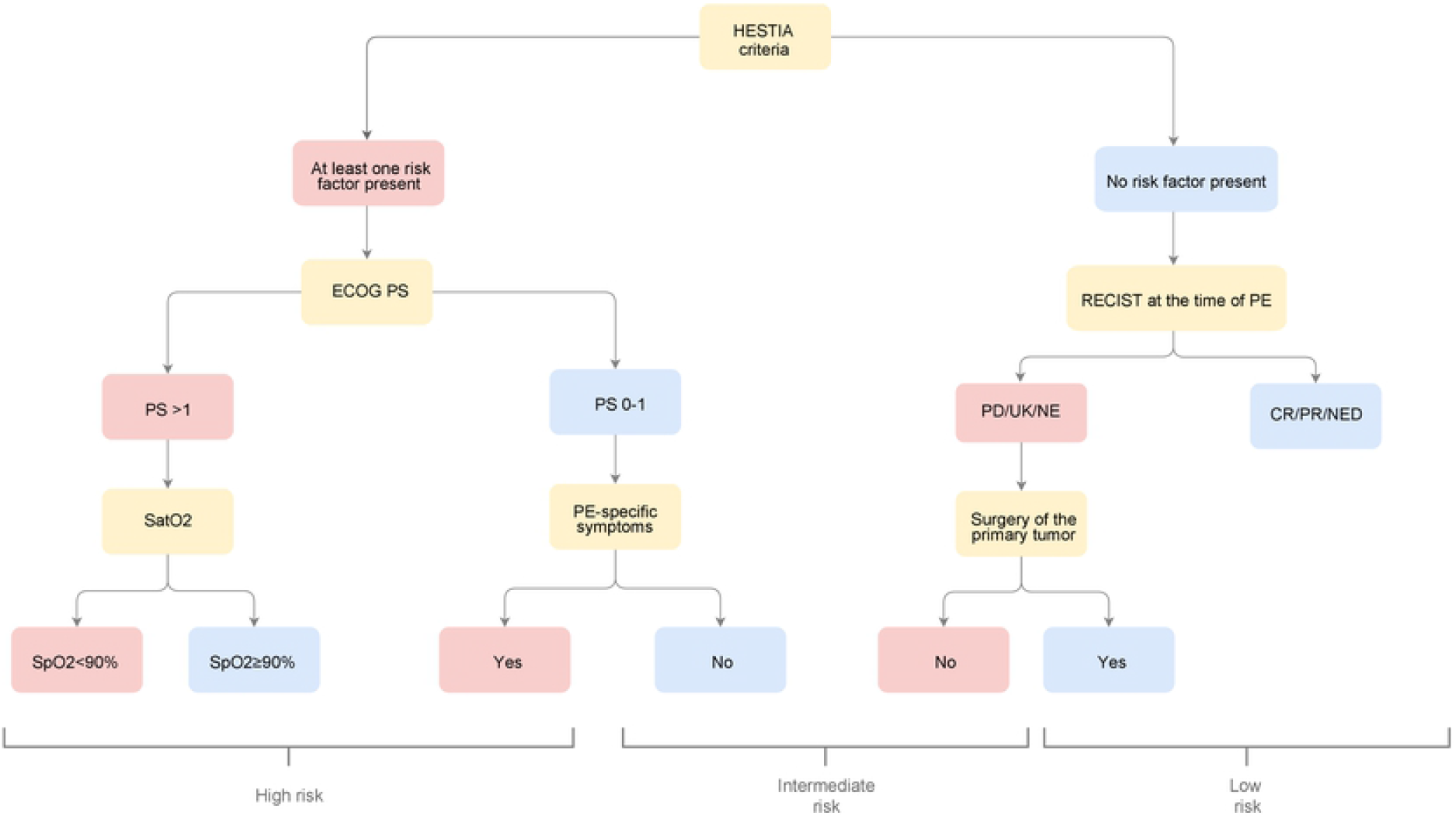
The EPIPHANY index. Abbreviations: ECOG PS, Eastern Cooperative Group Performance status; SpO2, oxygen saturation; PE, pulmonary embolism; RECIST, Response Evaluation Criteria In Solid Tumors; PD, progressive disease; UK, unknown; NE, not evaluable, CR, complete response; PR, partial response. The HESTIA criteria are typified by the presence of at least one of the following: systolic blood pressure <100 mm Hg, arterial oxygen saturation <90%, respiratory rate ≥30 breaths per minute, pulse ≥110 beats per minute, sudden or progressive dyspnea, other serious complications, constituting admission criteria in and of themselves, and clinically relevant bleeding, high risk of bleeding, or platelets <50 000 mm−3. These criteria should be assessed immediately prior to the time of radiological diagnosis of PE.

### Statistics

The estimation of relative frequency ***θ*** of complications based on the EPIPHANY Index categories was performed by means of the Bayesian alternative for the binomial test, applying Jeffreys non-informative prior [31]. The sample size was calculated to estimate ***θ*** in the low-risk EPIPHANY category with a desired degree of precision in the posterior distribution (***θ***=0.02; 95% HDI width <0.02 at least 90% of the time) [31]. Assuming approximately one-third low-risk subjects, a sample size of 900 participants was estimated. As measures of performance, sensitivity, specificity, positive and negative predictive values (PPV and NPV), and positive and negative likelihood ratios (PLR and NLR) were estimated. Causal inference as to the contribution of the individual variables was explored via frequentist multivariable logistic regression, including the 6 variables of the EPIPHANY index. Discrimination was appraised by comparing the area under the curve-receiver operating characteristics curve (AUC-ROC) using the DeLong test. OS was estimated using the Kaplan-Meier method. In the presence of competing risks, the cumulative incidence of rethrombosis and bleeding events was probed with the Aalen-Johansen estimator. The time to event functions were compared using Gray or log-rank tests in each case. Analyses were carried out using the R v4.05 software including the cmprsk, survival, pROC, and rjags libraries [32–35].

## RESULTS

### Patients

The sample consists of 900 patients treated between October 2017 and March 2020. Baseline characteristics can be found in **Table 1**. The median age was 66 years (range 21-94), with a predominance of males (57.4%, n=517), and good functional status (ECOG-PS 0-1, 64.6%, n=582). The most frequent tumors were broncho-pulmonary (26.3%, n=237), colorectal (18.6%, n=168), and breast (8.1%, n=73); most were stage IV (78.3%, n=705). Seven hundred participants (77.7%) were in active treatment, the most frequent anticancer therapy being chemotherapy (60.6%, n=546). The primary tumor had not been resected in two thirds (66.5%, n=599). Almost half (48.6%, n=437) of the episodes were unsuspected, asymptomatic. Full-dose low-molecular-weight heparin was the treatment administered in 92.9% (n=836).

**Table 1.** Baseline characteristics of the sample. Abbreviations: COPD, Chronic Obstructive Pulmonary Disease; CT, computed tomography ECOG-PS, Eastern Cooperative Oncology Group; NED, No Evidence of Disease; PE, Pulmonary Embolism; RECIST, Response Evaluation Criteria In Solid Tumors.

| **Baseline characteristics** | **N=900 (100%)** |
| --- | --- |
| **Sex, male** | 517 (57.4) |
| **Age, median (range)** | 66 (21-94) |
| **ECOG-PS** |  |
| 0 | 136 (15.1) |
| 1 | 446 (49.6) |
| 2 | 229 (25.4) |
| 3 | 82 (9.1) |
| 4 | 7 (0.8) |
| **Most common tumors** |  |
| *Lung* | 237 (26.3) |
| *Colorectal* | 168 (18.7) |
| *Breast* | 73 (8.1) |
| *Pancreatic* | 64 (7.1) |
| *Bladder* | 47 (5.2) |
| *Stomach* | 45 (5.0) |
| *Ovarian* | 40 (4.4) |
| *Central nervous system* | 31 (3.4) |
| *Endometrial* | 25 (2.8) |
| **TNM classification, stage IV** | 705 (78.3) |
| **RECIST at PE diagnosis** |  |
| *Not evaluable* | 418 (46.4) |
| *Progression* | 204 (22.7) |
| *Stable disease* | 144 (16) |
| *Partial response* | 94 (10.4) |
| *Complete response/ NED* | 40 (4.4) |
| **Comorbidities** |  |
| *Chronic cardiovascular disease* | 90 (10.0) |
| *COPD* | 79 (8.8) |
| *Chronic kidney disease* | 34 (3.7) |
| *Chronic liver disease* | 15 (1.4) |
| *Major surgery in the previous 3 months* | 50 (5.6) |
| **Smoking status** |  |
| *Non-smoker* | 160 (17.8) |
| *Ex-smoker* | 31 (3.4) |
| *Active smoker* | 312 (34.7) |
| *Unknown* | 397 (44.1) |
| **Previous thrombosis** | 139 (15.5) |
| **Anticoagulation therapy at diagnosis of PE** | 110 (12.3) |
| **Anti-aggregation therapy at diagnosis of PE** | 71 (7.9) |
| **PE diagnostic technique** |  |
| *Angio-CT* | 301 (33.4) |
| *Ventilation/ perfusion scintigraphy* | 18 (2.0) |
| *CT for response assessment* | 437 (48.6) |
| *CT for other reasons* | 144 (16) |
| Type of PE |  |
| Unilateral | 502 (55.8) |
| Bilateral | 398 (44.2) |
| Central | 209 (23.2) |
| Peripheral | 333 (37.0) |
| Central and peripheral | 358 (39.8) |
| Suspected | 319 (35.4) |
| Unsuspected, symptomatic | 144 (16.0) |
| Unsuspected, asymptomatic | 437 (48.6) |
| Type of care |  |
| Outpatient | 351 (39.0%) |
| Inpatient | 540 (61.0%) |

### Outcomes

Serious complications within 15 days were confirmed in 107 patients (11.8%, 95% CI, 9.8-14.1). The median time from PE diagnosis until the appearance of said complications was 4 days (range, 0-15). **Table 2** comprises a list of the complications, the most common being acute respiratory insufficiency (6.6%, n=59), major bleeding (2.6%, n=23), and shock (2.3%, n=21).

**Table 2.** Severe 15-day complications grouped by treatment site. Abbreviations: CPR, cardiopulmonary resuscitation (CPR); ICU, Intensive Care Unit; NIV, non-invasive mechanical ventilation; OTI, orotracheal intubation; RV, right ventricle; SBP, Systolic Blood Pressure. *Multiple-response variable.

| Complication* | Patients in outpatient care<br>N=351 (100%) | Patients in inpatient care<br>N=549 (100%) | Total<br>N=900 (100%) |
| --- | --- | --- | --- |
| Respiratory insufficiency | 4 (1.1) | 55 (10) | 59 (6.6) |
| Major bleeding | 3 (0.9) | 20 (3.6) | 23 (2.6) |
| Hypotension<br>(SBP <90 mmHg) | 0 | 21 (3.8) | 21 (2.3) |
| Sepsis | 1 (0.3) | 18 (3.3) | 19 (2.1) |
| ICU admission | 0 | 18 (3.3) | 18 (2) |
| RV failure | 0 | 16 (2.9) | 16 (1.8) |
| Renal insufficiency | 0 | 11 (2%) | 11 (1.2) |
| NIV | 0 | 7 (1.3%) | 7 (0.8) |
| OTI | 0 | 6 (1.1%) | 6 (0.7) |
| Fibrinolysis | 0 | 5 (0.9%) | 5 (0.6) |
| Need for CPR maneuvers | 0 | 1 (0.2%) | 1 (0.1) |
| Death | 4 (1.1) | 61 (11.1) | 65 (7.2) |
| Other complications | 1 (0.3) | 17 (3.1%) | 18 (2) |
| Any complication | 9 (2.6) | 98 (17.9) | 107 (11.9) |

At the time of analysis, 434 events of death (48.2%) were detected after a median follow up in patients still alive of 11.2 months (95% CI, 10.2-12.2). Median OS was 10.0 months (95% CI, 7.9-11.9). The causes of 30-day mortality are listed in **Annex Table 1**, the most prevalent being tumor progression. No deaths were due to rethrombosis in the first 30 days following PE diagnosis, albeit 6.7% of the expiries during this period were attributable to major bleeding.

### Validation of the EPIPHANY Index

The reconstruction of the decision tree is displayed in **Annex Figure 1**. Serious complications at 15 days arose in 2.4% of the patients with episodes classified as low-risk (95% highest density interval [HDI], 0.8-4.6%), in 5.5% (95% HDI, 2.9-8.7) of those with moderate-risk episodes, and in 21.0% (95% HDI, 17.0-24.0) of subjects with high-risk episodes (posterior probability ***θ***_***moderate-risk***_> ***θ***_***low-risk***_=95.8%, posterior probability ***θ***_***high-risk***_> ***θ***_***moderate-risk***_ =100%) (**Table 3**).

**Table 3.** Distribution of risk classes and 15-day serious complications according to the EPIHANY index/ modified HESTIA criteria. Abbreviation: HDI, highest density interval, n= number with complications; N= number of patients.

| Category | Complications, n/N | Complications, % (95% HDI) | Mortality, n/N | Mortality, % (95% HDI) |
| --- | --- | --- | --- | --- |
| Low-risk Epiphany | 5/ 232 | 2.2 (0.8-4.6) | 1/232 | 0.5 (0.0-2.0) |
| Intermediate-risk Epiphany | 12/ 229 | 5.5 (2.9-8.7) | 7/229 | 3.1 (1.3-5.9) |
| High-risk Epiphany | 90/ 439 | 20.5 (16.9-24.4) | 57/439 | 13.0 (10.0-16.4) |
| Low-risk HESTIA | 8/ 350 | 2.3 (1.0-4.2) | 5/ 350 | 1.4 (0.5-3.1) |
| High-risk HESTIA | 99/ 550 | 18.0 (14.9-21.4) | 60/ 550 | 10.9 (8.5-13.7) |

The Bayesian alternative for the binomial test is shown in **Figure 2**. The EPIPHANY Index correlated consistently with OS (**Figure 3**). Applying only the Hestia criteria, the 15-day rate of serious complications was 18% (95% HDI, 15-21%) in the presence of at least one factor vs 2.5% (95% HDI, 1.0-4.2) in the absence of all criteria (posterior probability **θ+**_**Hestia**_>**θ-**_**Hestia**_ =100%).

**Figure 2.**
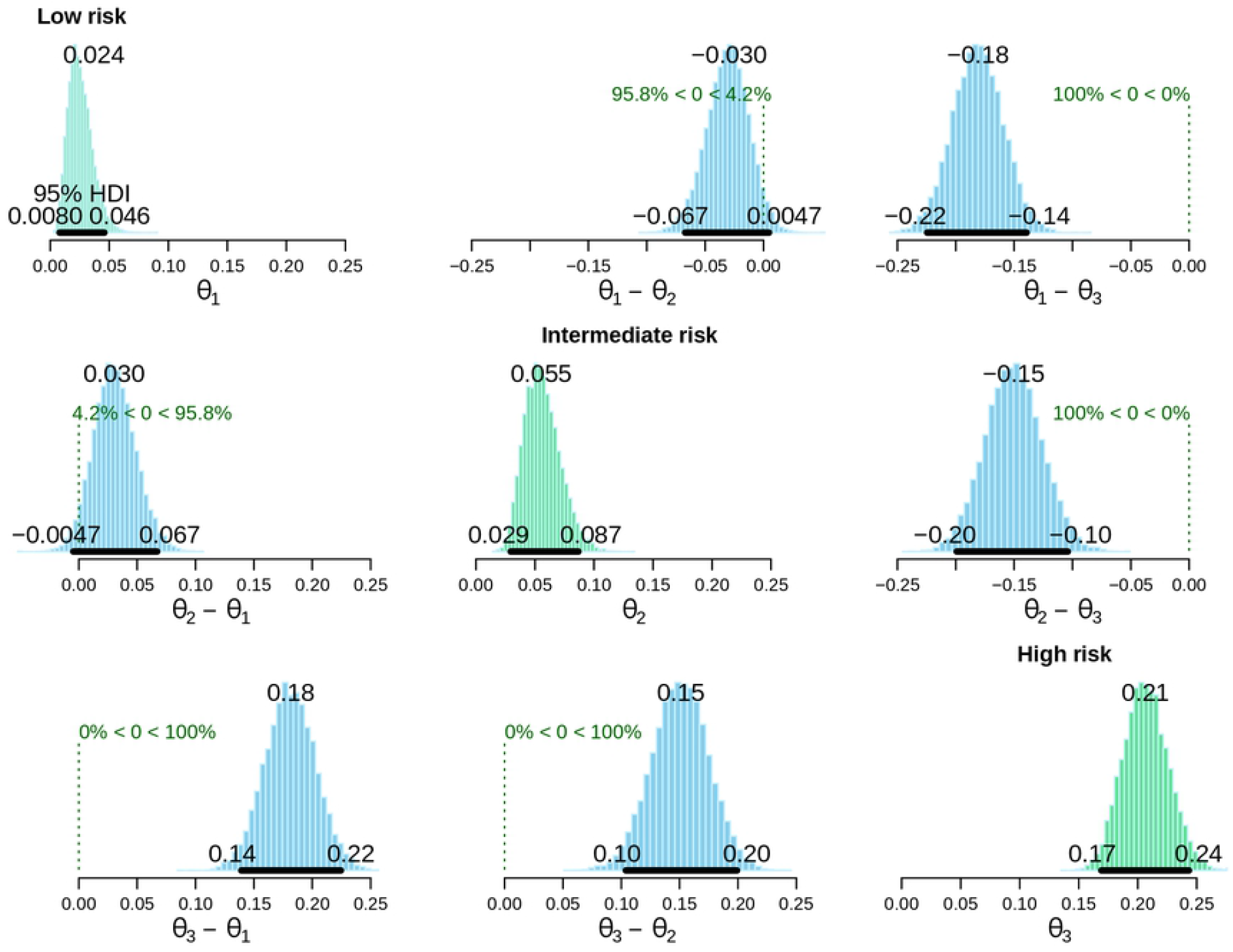
Bayesian alternative for the binomial test. The endpoint is the 15-day complication rate, represented by the θ parameter, the subscript representing the risk category (θ_1_ low risk, θ_2_ intermediate risk, θ_3_ high risk). The green panels characterize the posterior distribution of θ for each category. The blue panels display the posterior for the difference in proportions. In each case, the probability that the difference is greater or less than zero is reported.

**Figure 3.**
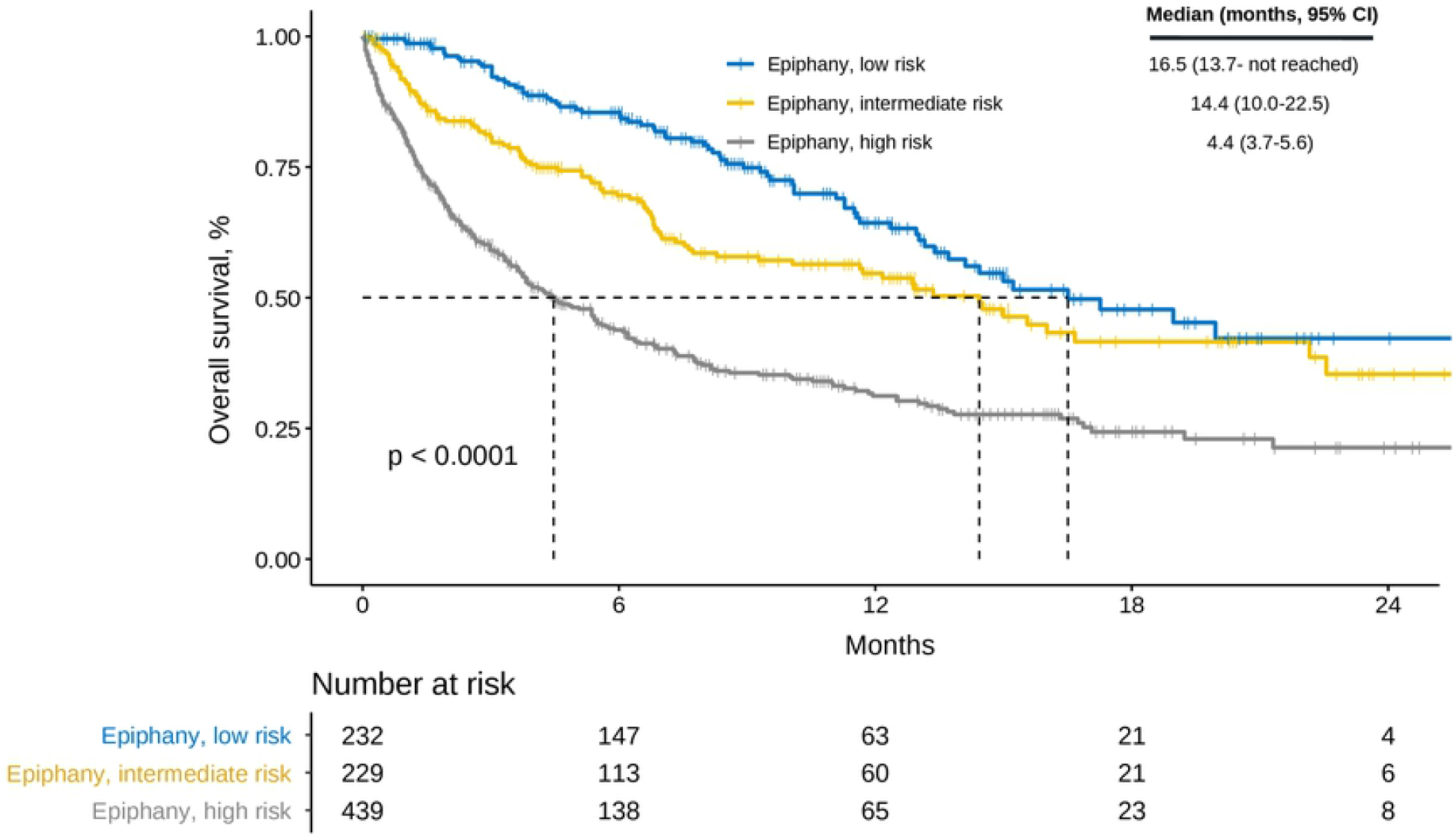
Overall survival based on the EPIPHANY index categories. Abbreviations: CI= confidence interval.

The dichotomized EPIPHANY Index (high vs other risks) is associated with a NPV of 97.8% (95% CI, 95.0-99.2) and NLR of 0.16 (95% CI, 0.07-0.39) (**Annex Table 2**). In episodes of suspected PE, the EPIPHANY Index is associated with a NPV of 90.9% (95% CI, 62.2-98.3) and NLR of 0.38. As for 30-day mortality, the EPIPHANY Index exhibits a NPV of 99.1% (95% CI, 96.9-99.8) and NLR of 0.06 (95% CI, 0.01-0.26) (**Annex Table 3**). Against other scores, the EPIPHANY index has a greater NPV to rule out complications in symptomatic PE. Both exhibited good discriminatory capacity in unsuspected PE (**Table 3**).

Different data splits based on geographic distribution did not reveal substantial changes with respect to overall classification performance (**Annex Figure 2**). The cumulative incidence of venous rethrombosis at 6 months as per the EPIPHANY Index was 2.9% (95% CI, 0.6-5.3) in low/moderate-risk vs 7.0% (95% CI, 4.9-9.1) in high-risk episodes (p-value=0.651, Gray’s test) (**Annex Figure 3**). The incidence of bleeding (any grade) at 6 months was 6.2% (95% CI, 2.9-9.5) in low/moderate-risk vs 12.7% (95% CI, 10.1-15.4) in high-risk episodes (Gray’s test, p-value=0.037) (**Annex Figure 4**).

In the logistic regression, the covariates having the greatest weight were the modified Hestia criteria, odds ratio (OR) 3.6 (95% CI, 1.5-8.4), and disease in progression or not evaluable, OR of 2.1 (95% CI, 1.1-4.0) (**Annex Table 4**).

### Performance in ambulatory patients

The 15-day rate of serious complications in ambulatory patients was 2.5% (95% HDI, 1.1-4.8). These complications were detected later in outpatients vs inpatients (median 9 vs 3 days, Wilcoxon’s test, p-value=0.025). None of the subjects managed in their homes died as a result of acute PE complications. Complications at 15 days were recorded in 5/273 subjects with low/intermediate EPIPHANY risk (2.1%, 95% HDI, 0.7-4.0) vs 4/78 (5.9%, 95% HDI, 1.7-11.9) in high EPIPHANY risk (posterior probability ***θ***_***high***_>***θ***_***moderate/low***_ =94.9%) (**Annex Table 5** and **Annex Figure 5**). **Annex Table 6** reflects the 30-day mortality rates. 4 patients (1.1%) treated as outpatients died during this period (2 cases were attributed to progression, 2 to mixed cause). The distribution of the modified Hestia criteria in ambulatory patients is displayed in **Annex Table 7**.

## DISCUSSION

This study has validated the EPIHANY Index, a new tool to predict serious complications in patients with PE and cancer, with potential usefulness for outpatient management decisions. This index is an extension of the Hestia criteria, developed to increase its discriminatory power in the oncological population by including specific variables, such as ECOG-PS, tumor response as per RECIST, surgery on the primary tumor, or symptoms [2,8,28]. The Hestia criteria have been used here in an adapted form to capture the higher risk of bleeding in this population [4,15] Our study confirms the validity of these criteria and their extension in the EPIPHANY Index, which endorses the use of sets of exclusion criteria similar to those proposed by other authors over sum scores [2,17,27]. Both are more reliable than other scales to rule out complications, inasmuch as they are capable of discriminating between incidental PEs not classifiable by other models.

Certain nuances must be taken into account when interpreting these results. First, the EPIPHANY Index predicts the 15-day complication rate, since this endpoint was considered to be the most relevant when making decisions about ambulatory management [2]. In contrast, the other scores predict 30-day all-cause mortality. This latter endpoint involves the risk of incurring in clinical contradictions, such as overtreating high-risk patients receiving palliative care, given that most predicted events will be attributable to tumor progression with a marginal role of the PE [8]. Second, the probability that the outpatient actually cares about is the NPV, and this parameter is higher in the EPIPHANY Index and Hestia criteria vs the other models. In exchange, all the models exhibited a low PPV, especially in scenarios that are *a priori* low-risk, such as incidental PE. This limitation is shared by other scores used in support care, generally more useful to rule out serious complications than to identify them [36]. By way of example, high-risk subjects as per RIETE [22] had a 22% complication rate in this sample, similar to that seen with Hestia/EPIPHANY high-risk individuals. Undoubtedly, the asymmetric consequences of each error (e.g., malapropos scaling back vs intensifying therapy) must be factored into any treatment algorithm. The most nefarious result is that subjects develop complications after being mistakenly classified as low-risk. In contrast, if we persevere with the pragmatic approach, the low PPV poses a tolerable risk. In fact, hospitalization founded on Hestia criteria is hardly negligible in practical terms, even in the absence of a true basis of severity. In any case, it is possible to envision predictors with a greater PPV, albeit disconnected from a utilitarian orientation, rendering their practicality questionable. Third, the capacity to discriminate prognosis in incidental episodes is one of the most noteworthy contributions, since the rest of the scores were designed for acute, symptomatic PE [37] and do not contain the variables that make it possible to discriminate levels of risk within unsuspected PE. Our results are consistent with the retrospective validation by Ahn et al, focused on incidental PE, that reported a 15-day complication rate of 3.4, 8.9, and 23.8% for low-, intermediate-, and high-risk EPIPHANY episodes [28]. In return, the EPIPHANY Index/Hestia criteria perform worse in acute, symptomatic PE, as patients in this scenario are rarely classified as low risk. Nevertheless, the NLR of EPIPHANY index/Hestia criteria applied to symptomatic PE is comparable to the remaining validated scales, reflecting the fact that operationally, they perform on a par. Finally, the EPIPHANY Index can discriminate bleeding risk, making it appealing in the oncologic population, as it captures multiple variables associated with bleeding that is the most serious iatrogenic complication.

Our study has several limitations. The most evident one is that its observational design did not set out to contemplate any intervention based on EPIPHANY and, in fact, the investigators were not explicitly aware of the classification. Yet, the subjects receiving ambulatory treatment in this series tended to *de facto* uphold most of the Hestia criteria, the most tolerated factors being the risk of bleeding (16%) and dyspnea (13%). Validation in clinical trials is needed. In any case, the favorable results of this empirical selection are similar to those reported with the most refined models [15,16]. The absence of a concordance analysis and the unblinded evaluation of outcomes are other limitations [38]. Technically, another limitation is the frailty of the classifiers based on decision trees, with interactions that are often not validated or vary across populations. This method was applied to adapt to decision-making algorithms in the real world [8]. It is certainly feasible that other, more complex methods based on ensembles (e.g., random forest, gradient boosting), might improve the PPV. Another limitation is that only Spaniards have been included. Caution is advised when extrapolating to other populations, although, a priori, we would not expect large differences to emerge.

Ultimately, in this prospective, multicenter study, we have validated the EPIPHANY Index/Hestia criteria in patients with acute, cancer-related PE. This can help to pave the way to standardize decision-making in a scenario in which there was no quality evidence. Our results are relevant to supporting ambulatory management of patients with low-risk PE, particularly in the case of unsuspected events, inasmuch as they already have a lower risk of complications.

## Data Availability

All relevant data are within the manuscript and its Supporting Information files.

## Ethics approval

The study was approved by the Research Ethics Committee of the Hospital General Universitario José María Morales Meseguer (code: C.P. PERSEO - C.I. EST: 57/17, 26 October 2017) and by the Spanish Agency of Medicines and Medical Devices (AEMPS) (6 October 2017). The study has been conducted in accordance with the ethical standards of the 1964 Declaration of Helsinki and its later amendments. This study is an observational, non-interventionist trial.

## Consent to participate

Signed informed consent was obtained from all participants.

## Consent for publication

Informed consent and approval by the competent national authorities includes permission for publication and dissemination of the data.

## Authors’ contributions

M.S.C., A.C.B., and P.J.F. developed the project, analyzed the data, and drafted the manuscript.

The other authors recruited patients and provided clinical information, comments, and improvements to the manuscript. All authors participated in the interpretation and discussion of the data, as well as the critical review of the manuscript.

---

**Annex Figure 1.**
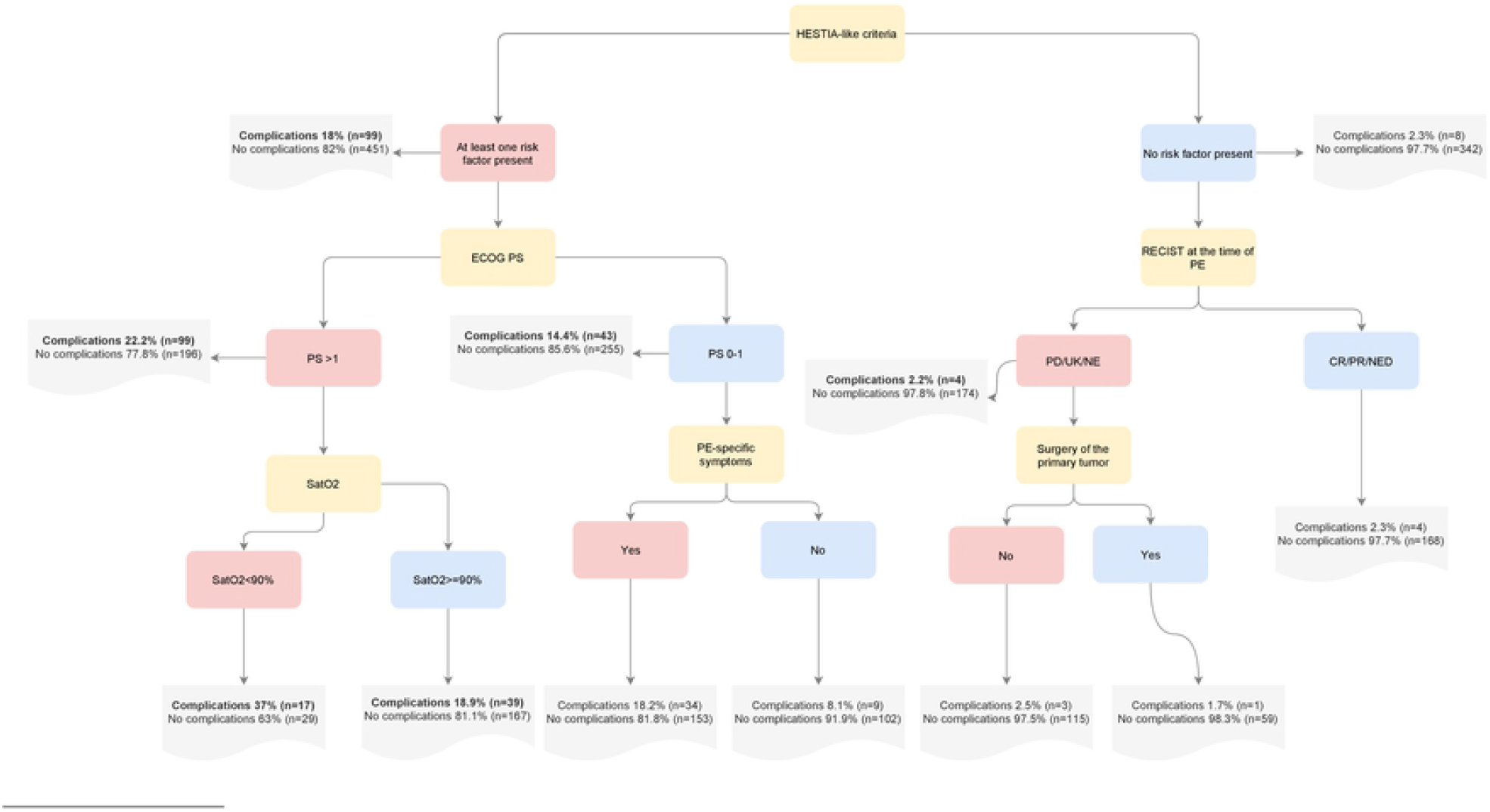
Reconstruction of the EPIPHANY Index with the PERSEO Study data. Abbreviations: ECOG PS= Eastern Cooperrative Group Performance Status, PE, Pulmonary embolishm, PD= progressive disease, UK= unknown, NE=not evaluated, CR= complete response, PR= partial response, NED= no evidence of disease

**Annex Figure 2.**
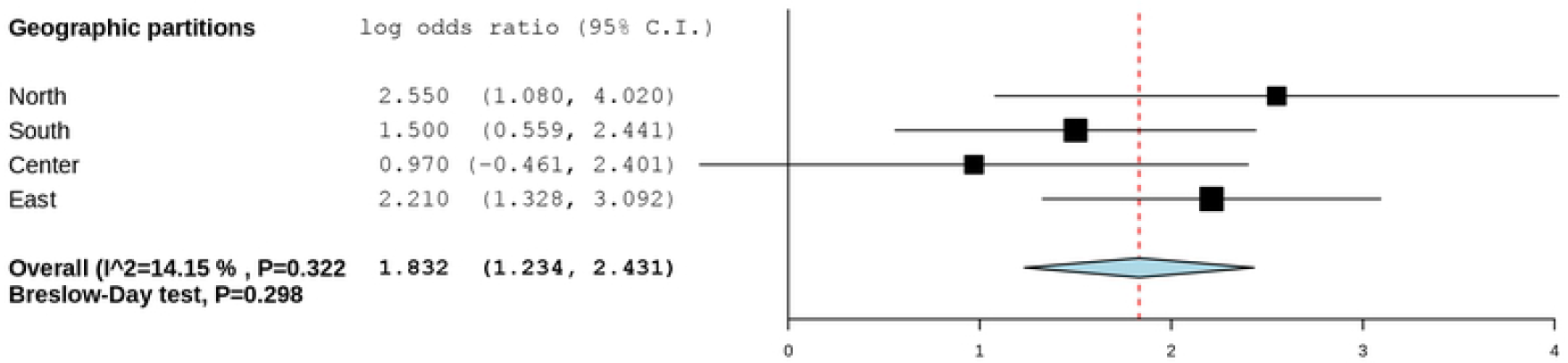
Discrimination capacity of the EPIPHANY index in geographic partitions. Note: The index has been dichotomized (high risk vs.Others). CI= Confidence Interval

**Annex Figure 3.**
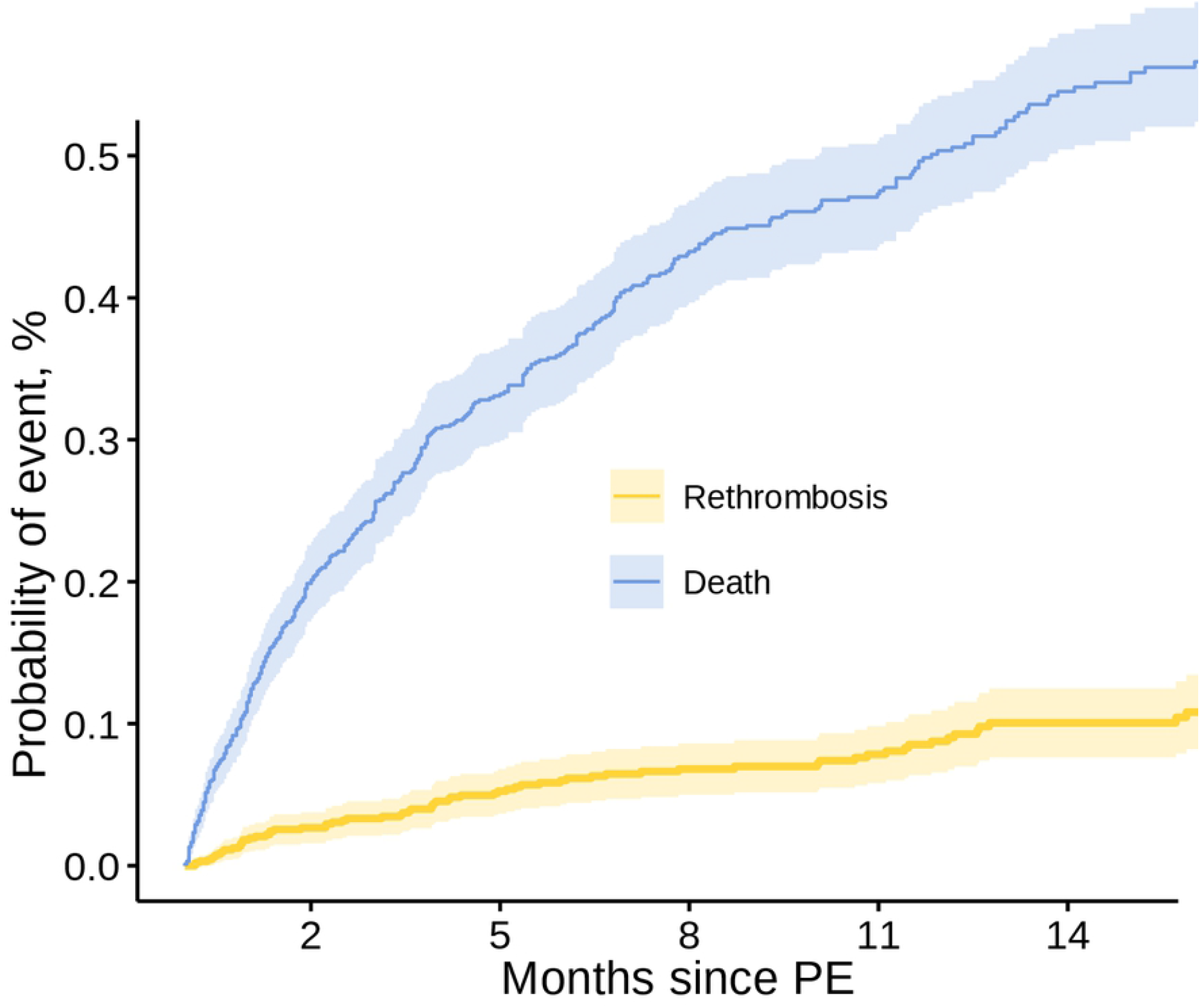
Rethrombosis and death.

**Annex Figure 4.**
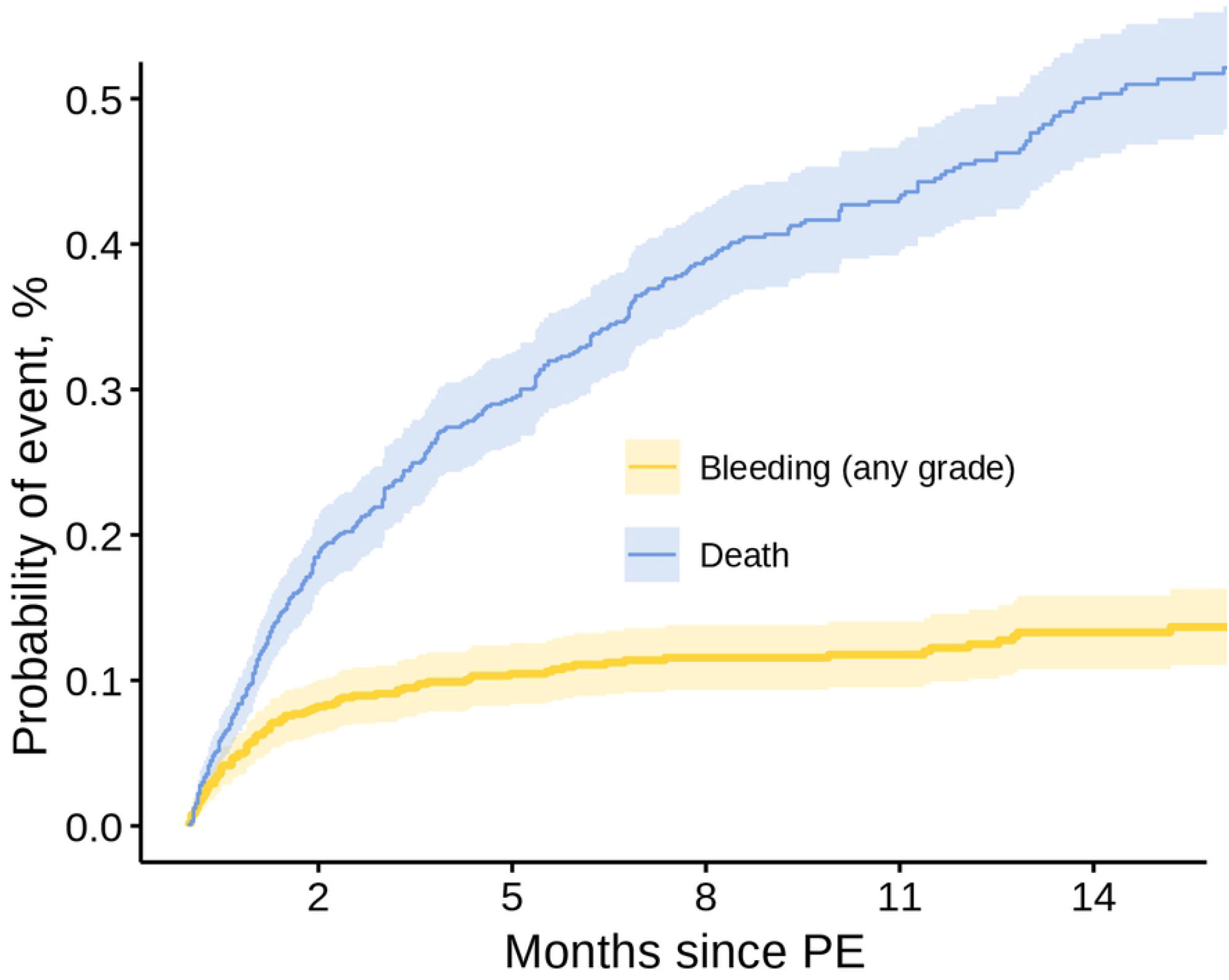
Bleeding and death.

**Annex Figure 5.**
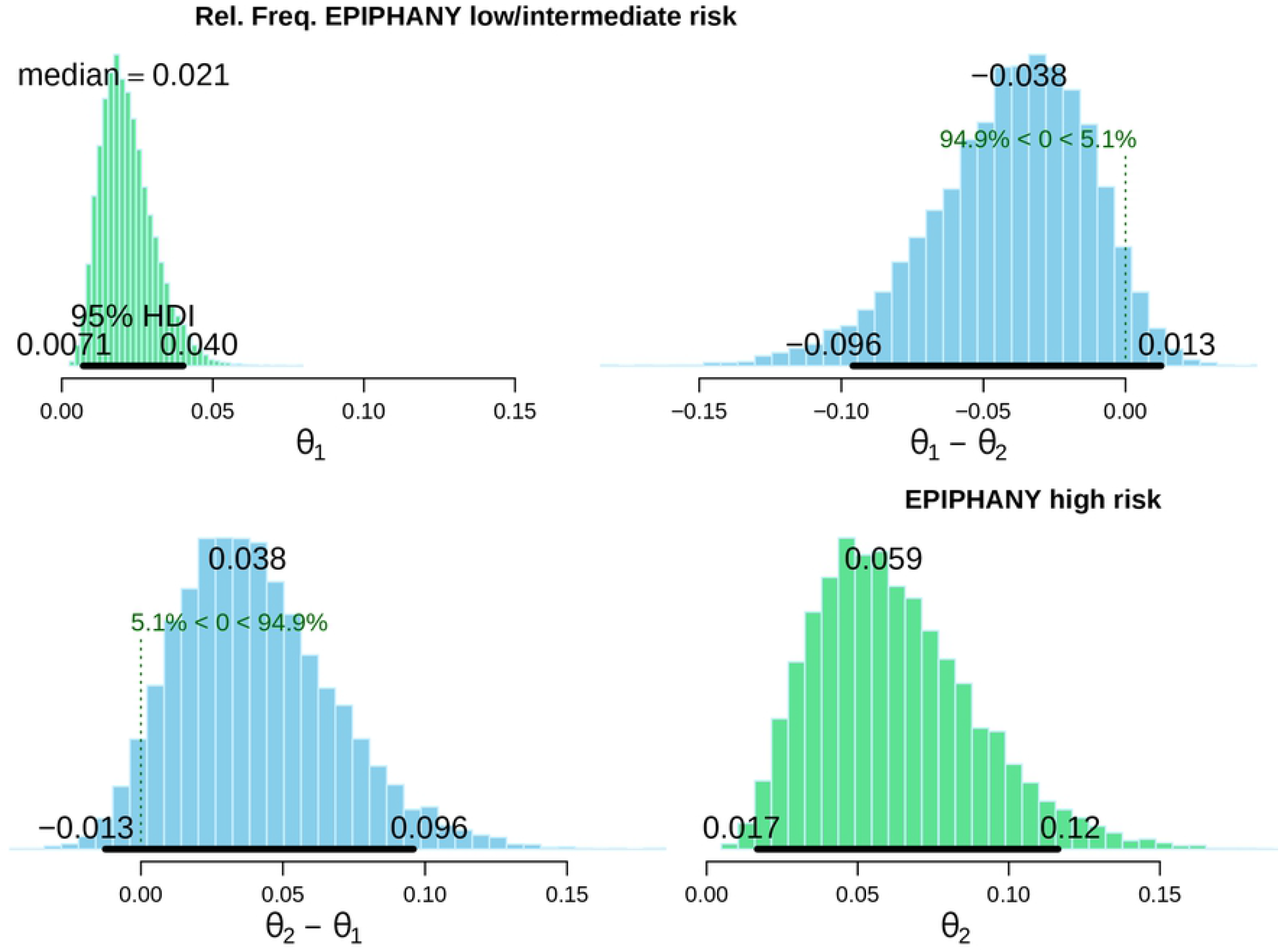
Serious Complications at 15 days in patients treated at home.

## REFERENCES

1. Monreal M, Falgá C, Valdés M, Suárez C, Gabriel F, Tolosa C, et al. Fatal pulmonary embolism and fatal bleeding in cancer patients with venous thromboembolism: findings from the RIETE registry. J Thromb Haemost. 2006;4: 1950–6. doi:10.1111/j.1538-7836.2006.02082.x

2. Carmona-Bayonas A, Font C, Jiménez-Fonseca P, Fenoy F, Otero R, Beato C, et al. On the necessity of new decision-making methods for cancer-associated, symptomatic, pulmonary embolism. Thromb Res. 2016;143. doi:10.1016/j.thromres.2016.05.010

3. Safriel Y, Zinn H. CT pulmonary angiography in the detection of pulmonary emboli: a meta-analysis of sensitivities and specificities. Clin Imaging. Elsevier; 2002;26: 101–105.

4. Carmona-Bayonas A, Gómez D, de Castro EM, Segura PP, Langa JM, Jimenez-Fonseca P, et al. A snapshot of cancer-associated thromboembolic disease in 2018–2019: First data from the TESEO prospective registry. Eur J Intern Med. Elsevier; 2020;78: 41–49.

5. Lehnert P, Lange T, Møller CH, Olsen PS, Carlsen J. Acute pulmonary embolism in a national Danish cohort: increasing incidence and decreasing mortality. Thromb Haemost. Schattauer GmbH; 2018;118: 539–546.

6. Meyer H-J, Wienke A, Surov A. Incidental pulmonary embolism in oncologic patients—a systematic review and meta-analysis. Support Care Cancer. Springer; 2021;29: 1293–1302.

7. Dentali F, Ageno W, Becattini C, Galli L, Gianni M, Riva N, et al. Prevalence and clinical history of incidental, asymptomatic pulmonary embolism: a meta-analysis. Thromb Res. Elsevier; 2010;125: 518–522.

8. Carmona-Bayonas A, Jiménez-Fonseca P, Font C, Fenoy F, Otero R, Beato C, et al. Predicting serious complications in patients with cancer and pulmonary embolism using decision tree modelling: The EPIPHANY Index. Br J Cancer. 2017;116: 994–1001. doi:10.1038/bjc.2017.48

9. Svendsen E, Karwinski B. Prevalence of pulmonary embolism at necropsy in patients with cancer. J Clin Pathol. BMJ Publishing Group Ltd and Association of Clinical Pathologists; 1989;42: 805–809.

10. Hendriks S V, Huisman M V, Eikenboom JCJ, Fogteloo J, Gelderblom H, van der Meer FJM, et al. Home treatment of patients with cancer-associated venous thromboembolism–An evaluation of daily practice. Thromb Res. Elsevier; 2019;184: 122–128.

11. Font C, Carmona-Bayonas A, Beato C, Reig Ò, Sáez A, Jiménez-Fonseca P, et al. Clinical features and short-term outcomes of cancer patients with suspected and unsuspected pulmonary embolism: the EPIPHANY study. Eur Respir J. Eur Respiratory Soc; 2016; 1600282.

12. Font C, Carmona-Bayonas A, Fernández-Martinez A, Beato C, Vargas A, Gascon P, et al. Outpatient management of pulmonary embolism in cancer: data on a prospective cohort of 138 consecutive patients. J Natl Compr Canc Netw. 2014;12.

13. Rubio-Salvador AR, Escudero-Vilaplana V, Rodríguez JAM, Mangues-Bafalluy I, Ferrán BB, Collado CG, et al. Cost of Venous Thromboembolic Disease in Patients with Lung Cancer: Costecat Study. Int J Environ Res Public Health. Multidisciplinary Digital Publishing Institute; 2021;18: 394.

14. Squizzato A, Galli M, Dentali F, Ageno W. Outpatient treatment and early discharge of symptomatic pulmonary embolism: a systematic review. Eur Respir J. 2009;33: 1148–55. doi:10.1183/09031936.00133608

15. Zondag W, Mos ICM, Creemers-Schild D, Hoogerbrugge a DM, Dekkers OM, Dolsma J, et al. Outpatient treatment in patients with acute pulmonary embolism: the Hestia Study. J Thromb Haemost. 2011;9: 1500–1507. doi:10.1111/j.1538-7836.2011.04388.x

16. Siragusa S, Arcara C, Malato A, Anastasio R, Valerio MR, Fulfaro F, et al. Home therapy for deep vein thrombosis and pulmonary embolism in cancer patients. Ann Oncol. 2005;16 Suppl 4: iv136–v139. doi:10.1093/annonc/mdi923

17. Weeda ER, Kohn CG, Peacock WF, Fermann GJ, Crivera C, Schein JR, et al. External Validation of the Hestia Criteria for Identifying Acute Pulmonary Embolism Patients at Low Risk of Early Mortality. Clin Appl Thromb. SAGE Publications; 2016; 1076029616651147.

18. Prandoni P, Lensing AW a, Piccioli A, Bernardi E, Simioni P, Girolami B, et al. Recurrent venous thromboembolism and bleeding complications during anticoagulant treatment in patients with cancer and venous thrombosis. Blood. 2002;100: 3484–3488. doi:10.1182/blood-2002-01-0108

19. Jiménez D, Aujesky D, Moores L, Gómez V, Lobo JL, Uresandi F, et al. Simplification of the pulmonary embolism severity index for prognostication in patients with acute symptomatic pulmonary embolism. Arch Intern Med. 2010;170: 1383–1389. doi:10.1001/archinternmed.2010.199

20. Martín AJM, Zamorano MCR, Benéitez Mcv, Morán LO, Pérez ÁG, Jiménez MM. Outpatient management of incidental pulmonary embolism in cancer patient. Clin Transl Oncol. Springer; 2019; 1–4.

21. Kline JA, Roy PM, Than MP, Hernandez J, Courtney DM, Jones AE, et al. Derivation and validation of a multivariate model to predict mortality from pulmonary embolism with cancer: The POMPE-C tool. Thromb Res. 2012;129. doi:10.1016/j.thromres.2012.03.015

22. Den Exter PLPL, Gómez V, Jiménez D, Trujillo-Santos J, Muriel A, Huisman M V, et al. A clinical prognostic model for the identification of low-risk patients with acute symptomatic pulmonary embolism and active cancer. Chest. 2013;143: 138–145. doi:10.1378/chest.12-0964

23. Aujesky D, Obrosky DS, Stone RA, Auble TE, Perrier A, Cornuz J, et al. Derivation and validation of a prognostic model for pulmonary embolism. Am J Respir Crit Care Med. 2005;172: 1041–1046. doi:10.1164/rccm.200506-862OC

24. Wicki J, Perrier A, Perneger TV V., Bounameaux H, Junod AF. Predicting adverse outcome in patients with acute pulmonary embolism: A risk score. Thromb Haemost. 2000;84: 548–552.

25. Uresandi F, Otero R, Cayuela A, Cabezudo MA, Jimenez D, Laserna E, et al. [A clinical prediction rule for identifying short-term risk of adverse events in patients with pulmonary thromboembolism]. Arch Bronconeumol. 2007;43: 617–622.

26. Jiménez-Fonseca P, Carmona-Bayonas A, Martín-Pérez E, Crespo G, Serrano R, Llanos M, et al. Health-related quality of life in well-differentiated metastatic gastroenteropancreatic neuroendocrine tumors. Cancer Metastasis Rev. 2015;34. doi:10.1007/s10555-015-9573-1

27. Nguyen E, Caranfa J, Lyman GH, Kuderer NM, Stirbis C, Wysocki M, et al. Clinical Prediction Rules for Mortality in Patients with Pulmonary Embolism and Cancer to Guide Outpatient Management: a Meta-Analysis. J Thromb Haemost. 2017; doi:10.1111/jth.13921

28. Ahn S, Cooksley T, Banala S, Buffardi L, Rice TW. Validation of the EPIPHANY index for predicting risk of serious complications in cancer patients with incidental pulmonary embolism. Support Care Cancer. Springer; 2018;26: 3601–3607.

29. Sciences C for IO of M. International guidelines for ethical review of epidemiological studies. CIOMS, Geneva, CH; 1991.

30. Carmona-Bayonas A, Jiménez-Fonseca P, Virizuela Echaburu J, Antonio M, Font C, Biosca M, et al. Prediction of serious complications in patients with seemingly stable febrile neutropenia: validation of the Clinical Index of Stable Febrile Neutropenia in a prospective cohort of patients from the FINITE study. J Clin Oncol. American Society of Clinical Oncology; 2015;33: 465–471. doi:10.1200/JCO.2014.57.2347

31. Kruschke J. Doing Bayesian data analysis: A tutorial with R, JAGS, and Stan. Academic Press; 2014.

32. Gray B. cmprsk: Subdistribution analysis of competing risks. URL http://CRANR. 2013;

33. Therneau TM, Lumley T. Package ‘survival’ [Internet]. 2016. Available: https://cran.r-project.org/package=survival

34. Plummer M, Stukalov A, Denwood M, Plummer MM. Package ‘rjags.’ Vienna, Austria. 2016;

35. Robin X, Turck N, Hainard A, Tiberti N, Lisacek F, Sanchez J-C, et al. pROC: an open-source package for R and S+ to analyze and compare ROC curves. BMC Bioinformatics. BioMed Central; 2011;12: 1–8.

36. Fonseca PJ, Carmona-Bayonas A, García IM, Marcos R, Castañón E, Antonio M, et al. A nomogram for predicting complications in patients with solid tumours and seemingly stable febrile neutropenia. Br J Cancer. Nature Publishing Group; 2016; 1191–1198.

37. van Es N, Di Nisio M, Cesarman G, Kleinjan A, Otten H-M, Mahé I, et al. Comparison of risk prediction scores for venous thromboembolism in cancer patients: a prospective cohort study. Haematologica. 2017

38. Breiman L. Random forests. Mach Learn. Springer; 2001;45: 5–32.

